# Causal associations between iron status and sepsis: a Mendelian randomisation analysis

**DOI:** 10.1101/2022.04.29.22274435

**Authors:** Fergus Hamilton, Ruth Mitchell, Haroon Ahmed, Peter Ghazal, Nic Timpson

## Abstract

Iron deficiency is associated with a substantial burden of morbidity. However, supplementation of iron has been linked to increased rates of serious infection in randomised trials of children in sub-Saharan Africa. Randomised trials in other settings have been inconclusive and it is unknown if changes in levels of iron biomarkers – a mark of setpoint changes in iron homeostasis - are linked to sepsis in these other settings. We used genetic variants associated with levels of iron biomarkers as instrumental variables in a Mendelian randomisation (MR) analysis to test the hypothesis that increasing levels of iron biomarkers increase the risk of sepsis. In observational and MR analyses we found that increases in iron biomarkers increase the risk of sepsis. In stratified analyses, we show that this risk may be larger in those with iron deficiency and/or anaemia. Taken together, results here suggest a required caution in supplementation of iron and underline the role of iron homeostasis in severe infection.

## Introduction

Iron is a critical element for many biological processes and potentially harmful effects occur with iron deficiency or overload. Globally, iron deficiency remains a major problem in both low to middle and high-income countries.^1^ Although iron supplementation for those with iron deficiency anaemia (IDA) is uncontroversial, uncertainty remains about the role of supplementation in other clinical scenarios, particularly in iron deficiency without anaemia (IDWA). Despite the lack of clear evidence of benefit in randomised trials, there is increasing use of intravenous and/or oral iron both on and off label; often for symptoms attributed to iron deficiency.^2–4^ However, it is also clear that iron excess can be harmful, with people who are genetically predisposed to high iron (e.g. patients with mutations in *HFE*) developing a complex and broad range of syndromes, including an increased risk of infection.^5^

There has been longstanding interest in iron levels and the risk and outcome of infection due to the critical role of iron as a nutrient for both fungal and bacterial pathogens and the balance between the host and pathogen in management of iron. Hepcidin is positively regulated by the inflammatory cytokine interleukin-6, resulting in rapid reduction in serum iron and increases in serum ferritin resulting in iron replete macrophages.^6^ The inflammatory regulated hepcidin-ferroportin axis is a common response to bacterial infection in humans.^7^ This response is thought to be an attempt to reduce available iron to invading pathogens and aligns with evidence from multiple laboratory studies that many common pathogens grow more readily in serum that is high in iron.^8^

Iron status is a complex trait and no single biomarker reliably represents total body iron state. Cellular location, redox state, and availability of iron are all relevant in understanding iron state. Markers used in clinical assessment of acute iron status, that rapidly (over hours) alter with iron supplementation or in response to physiological demands, include serum iron and the total iron binding capacity (TIBC), which, as a ratio (serum iron / TIBC), equates to the transferrin saturation (TSAT). TSAT levels clinically indicate the availability of iron for erythropoiesis and are low in iron deficiency and high in overload.^9^

In contrast, ferritin – an iron storage protein – is generally considered a biomarker of iron body stores in a non-inflammatory state. TSAT and ferritin are used as the main diagnostic tests for iron deficiency anaemia worldwide and are measured more commonly than the other biomarkers. The master regulator for systemic iron homeostasis is the liver derived hepcidin that inhibits the only iron transporter, ferroportin, in iron-absorptive gut enterocytes and iron-recycling macrophages.^7^

Although all four biomarkers are routine tests, ferritin is tested more commonly than the others as it is used as the initial diagnostic test for anaemia, and is used to stratify patients as iron deficient, with a traditional cut off of 50ug/L as defining iron deficiency, although it is recognised this is a continuum.^1^

The alteration of the iron homeostasis setpoint in infection and inflammation raises valid concerns about iron supplementation. The strongest evidence for the relationship between iron supplementation and adverse infection outcomes was found from a large randomised controlled trial of iron and zinc supplementation in children from Zanzibar.^10^ The trial was stopped early due to increased risks of severe illness and death in those assigned to the iron and zinc arm, with much of the increased risk attributed to malaria.

However, subsequent randomised trials have not identified this effect and the data remain controversial and inconsistent.^11^ In adult settings, a recent meta-analysis of all randomised controlled trials of intravenous iron found increased risks of infection in those randomised to receive intravenous iron, but there was substantial uncertainty around the effect size.^12^ Because of the limited number of studies, this meta-analysis was unable to compare oral iron vs no supplementation at all.

Uncertainty therefore remains around whether increased iron is associated with increased rates of sepsis. Mendelian randomisation (MR) – a form of applied genetic epidemiology which deploys instrumental variable analysis using germline genetic variants – has been used as an approach to estimate causal relationships pertinent to iron status.^13–15^ In this study, we set out to assess the relationship between iron status biomarkers and sepsis using a combination of observational and Mendelian randomisation analyses, in a large UK cohort study (UK Biobank).

## Main

### Observational data

#### Demographics and cohort definition

In our observational analysis, we used UK Biobank, a large adult volunteer cohort with linked primary and secondary care data that includes blood tests taken in primary care for routine reasons.^16^ We aimed to investigate whether higher levels of ferritin (the only iron biomarker routinely clinically tested, and so available for our analysis) were associated with increased rates of sepsis, using regression models adjusted for relevant covariates.

We identified 222,081 participants in UK Biobank with linked primary care and hospital admission data. Amongst this cohort, 195,444 ferritin tests were recorded in the primary care records of 82,678 participants. **Supplementary Figure 1** describes the flow through the study. The median ferritin level was 105 ug/L for men, and 68ug/L for women, while the distribution was approximately log-normal (**Supplementary Figure 2**). For our analysis population, we included those with ferritin levels taken after each participant was recruited to UK Biobank [2006-2014], leading to 72,865 participants. The tested cohort (those with a recorded ferritin test) differed from the cohort who were not tested, with more females (64% vs 53%) and with a different pattern of comorbidities (**Table 1)**, reflecting the differences in testing conditional on illness.

**Table 1:**
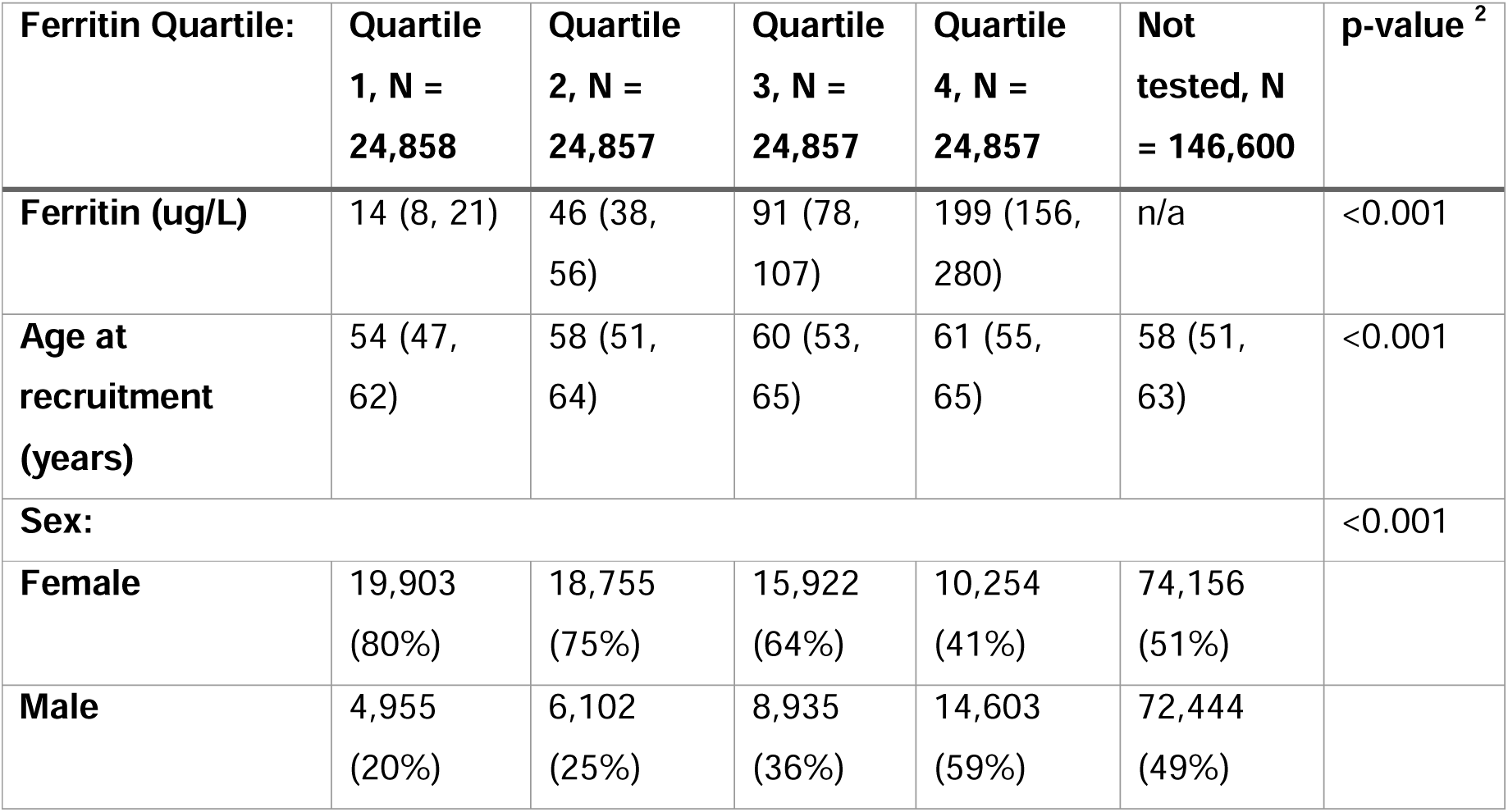
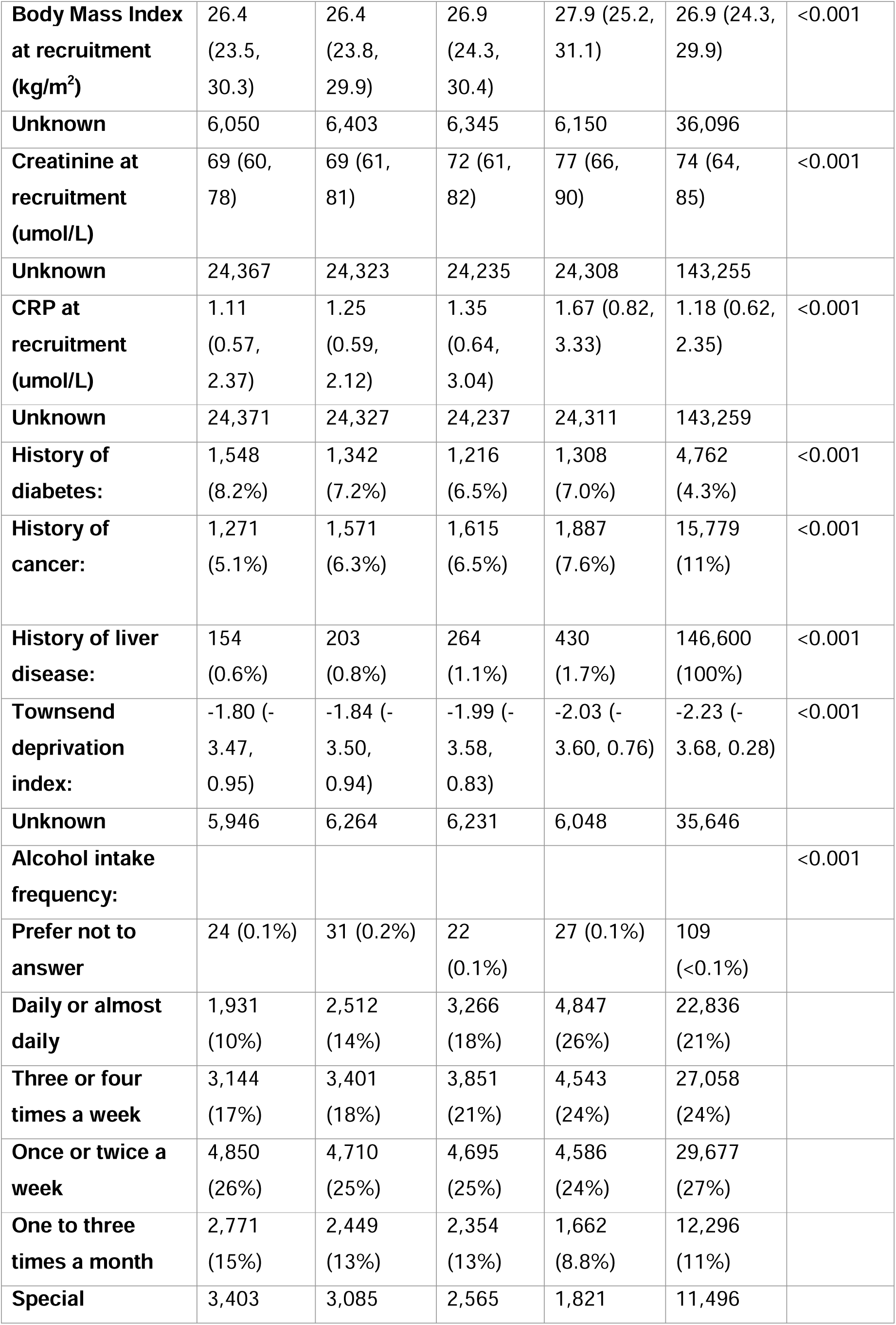
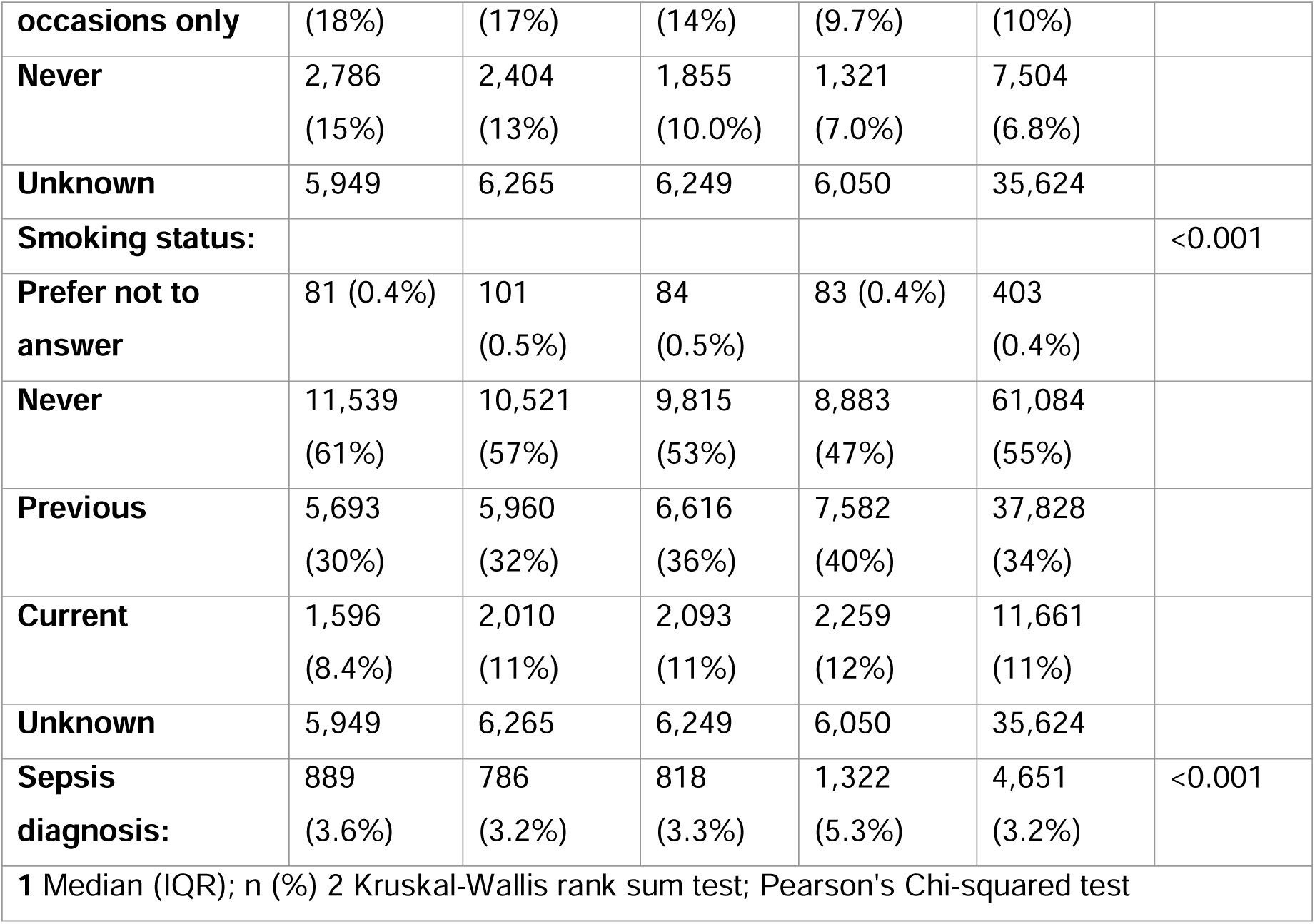
Associations between ferritin testing status and level on potential confounders.

We then used linked hospital admission data provided by UK Biobank (Hospital Episode Statistics data) to identify cases of sepsis. We used ICD-10 coding to identify admissions (codes A02, A39, A40, A41) in this cohort. There were 2,399 subsequent diagnoses of sepsis recorded in the tested cohort with a median time between ferritin result and sepsis diagnosis of 1,943 days (IQR 1,060 – 3,105).

#### Associations with sepsis

We went on to perform logistic regression to test the association between ferritin (untransformed) and subsequent sepsis diagnoses, adjusted for age, UK Biobank recruitment centre, smoking status, alcohol usage, deprivation and clinical comorbidities.

In this model increasing ferritin level was associated with an increased odds of sepsis (OR 1.05; 95% CI 1.04-1.06, p < 1.1 × 10^−11^ for every 100ug/L increase in ferritin) (**Supplementary Table 1 for full regression output**), with similar results in unadjusted models. We then went on to explore non-linearity, given that both low and high ferritin are associated with disease. In restricted cubic spline models (**Figure 1**) we found evidence for a U-shaped relationship in the early part of the ferritin distribution, with those at both extremes of ferritin levels at higher risk of sepsis, although the large bulk of the distribution lies within the linear association between increasing ferritin and increasing odds of sepsis.

**Figure 1:**
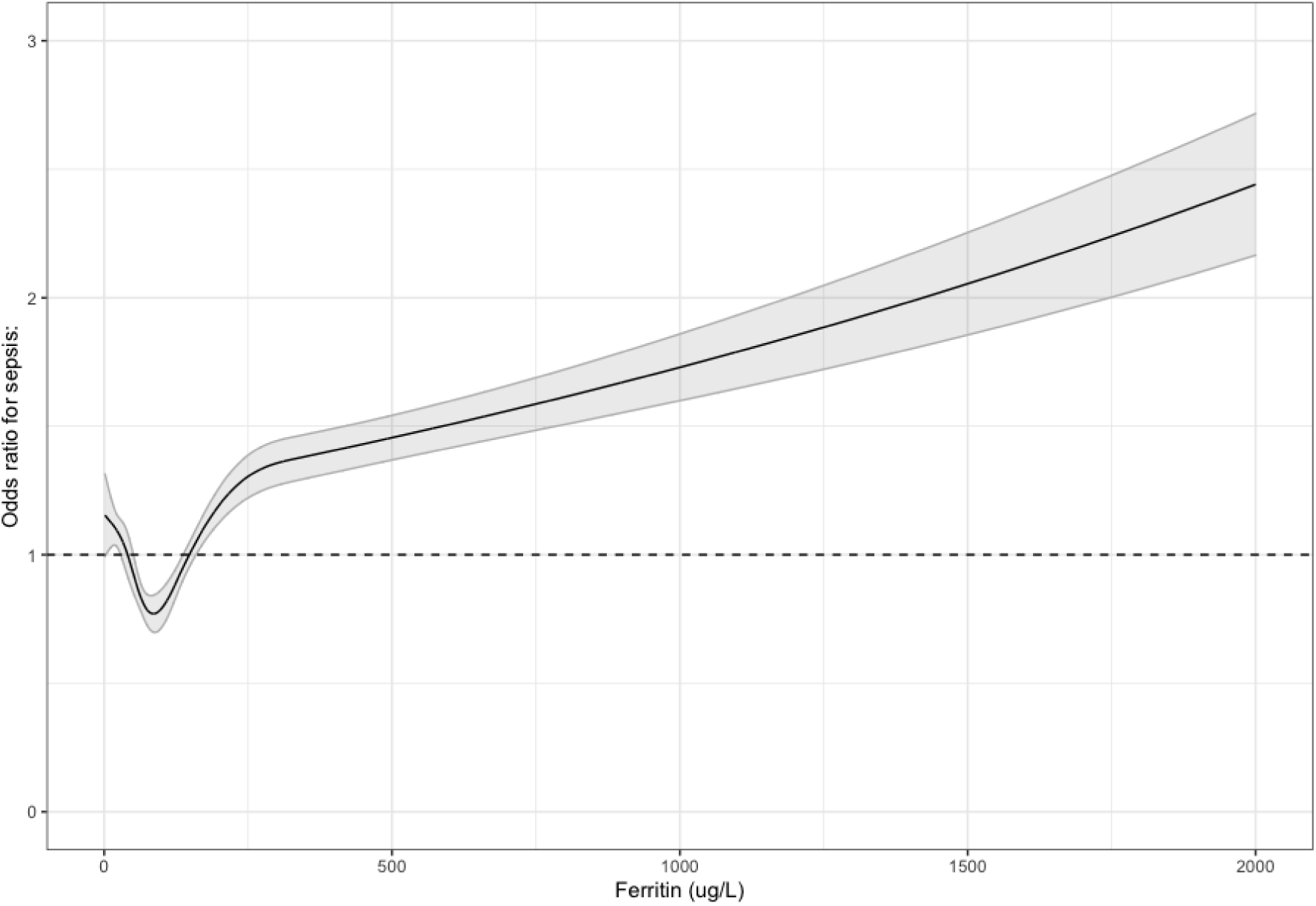
Restricted cubic spline plots of ferritin level and odds ratio for sepsis. The dashed line represents an odds ratio of 1, representing a null association.

As the management of iron and incidence and causes of iron deficiency vary by sex, we re-ran models stratified by sex. There were some differences in the magnitude of risk, with women having comparatively more risk with the same ferritin level. The odds of sepsis start to increase in women at around 68ug/L and in men at around 105ug/L (levels that are generally considered physiologically normal) and are around the centre of distribution of ferritin levels for this cohort (median Ferritin for men 108ug/L, for women 54ug/L). These results are shown in **Figure 2**, focussing on the range of 50-400 ug/L, with the full range visualised in **Supplementary Figure 3**.

**Figure 2:**
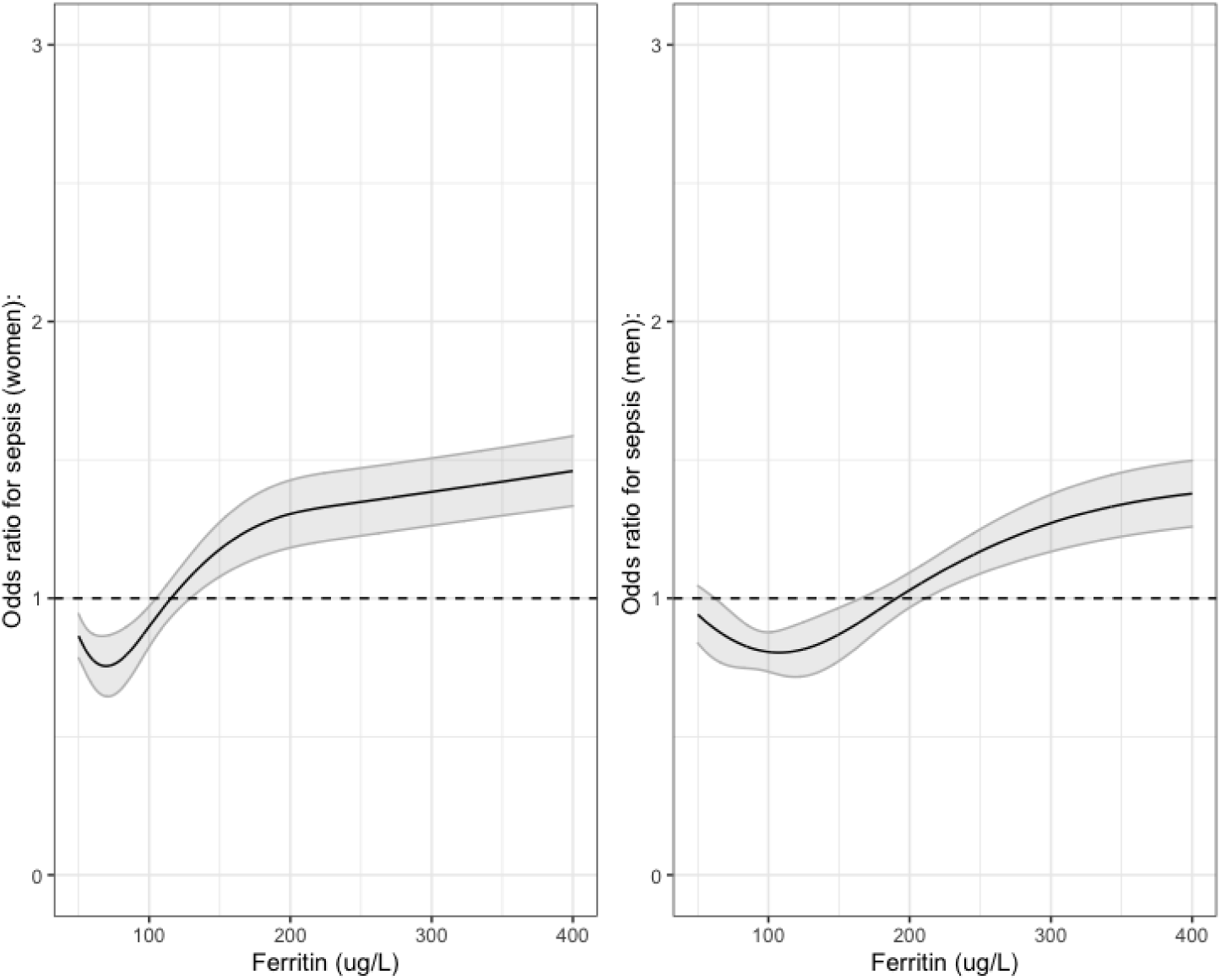
Odds ratios for sepsis with increasing ferritin between 50 – 450ug/L, with the dashed line representing an odds ratio of 1, a null association.

As increased rates of sepsis in those with extremely low ferritin was likely to be confounded by disease state (i.e. the disease leading to iron deficiency could lead to sepsis) we re-ran the linear analyses excluding all with ferritin results below 50ug/L. Effect estimates were similar, although larger, with an OR of 1.07 (95% CI 1.05-1.08, p = 5.8 × 10^−13^ per 100ug/L). However, interpretation of these estimates causally is difficult, due to conditioning by both disease state, demographics, and testing. Therefore, we went on to perform two sample Mendelian randomisation, to assess whether the association between iron status and sepsis was causal, and to generate estimates for comparison with iron supplementation.

#### Mendelian randomisation

##### Generation of instruments

Two sample Mendelian randomisation uses single nucleotide polymorphisms (SNPs), that are known to be associated with a trait (in this case iron biomarkers) in one study as proxy measures for this trait in a second study able to reassess associations with health outcome in an instrumental variable analysis.^15^

To generate instruments for ferritin, we extracted linkage disequilibrium independent (r2 <0.01), genome-wide significant (5 × 10^−8^) SNP’s from a recent meta-analysis of iron status, where each biomarker had been rank-based inverse normal transferred prior to analysis.^17^ Details of the cohort, genetic quality control, and methodology are in the methods.^18^ For serum ferritin we included 50 SNPs. Although we were unable to perform observational analyses for them, were also able to derive instruments for iron (23 SNPs), TSAT (17 SNPs), and (TIBC 26 SNPs). All included SNPS are available in **Supplement S4**.

##### Development of outcome GWAS

To generate outcome summary statistics for use in Mendelian randomisation analyses, we identified cases of sepsis using the same methodology as in the observational analysis, but in all participants in UK Biobank of European ancestry (case and ancestry definitions in the methods). In total, 12,664 cases were included, with 462,869 controls, giving a frequency of incident cases of 2.5%. We subsequently performed a case-control GWAS on quality controlled genetic data, with details of QC here^19^ and in the methods using regenie. regenie was chosen as it has better performance than traditional linear mixed models (e.g. BOLT-LMM) where there is a large case-control imbalance (as in this case).^20^ In total, we tested 12,224,3568 variants. For our main analysis, we only included participants with sepsis before the age of 75, as genetic influences on sepsis are likely to wane at the extremes of age. Manhattan and quantile-quantile plots are shown in **Supplementary Figures 4** and **5. Figure 3** describes the instruments and outcomes used for this analysis, with detailed description of the included cohorts in the methods.

**Figure 3:**
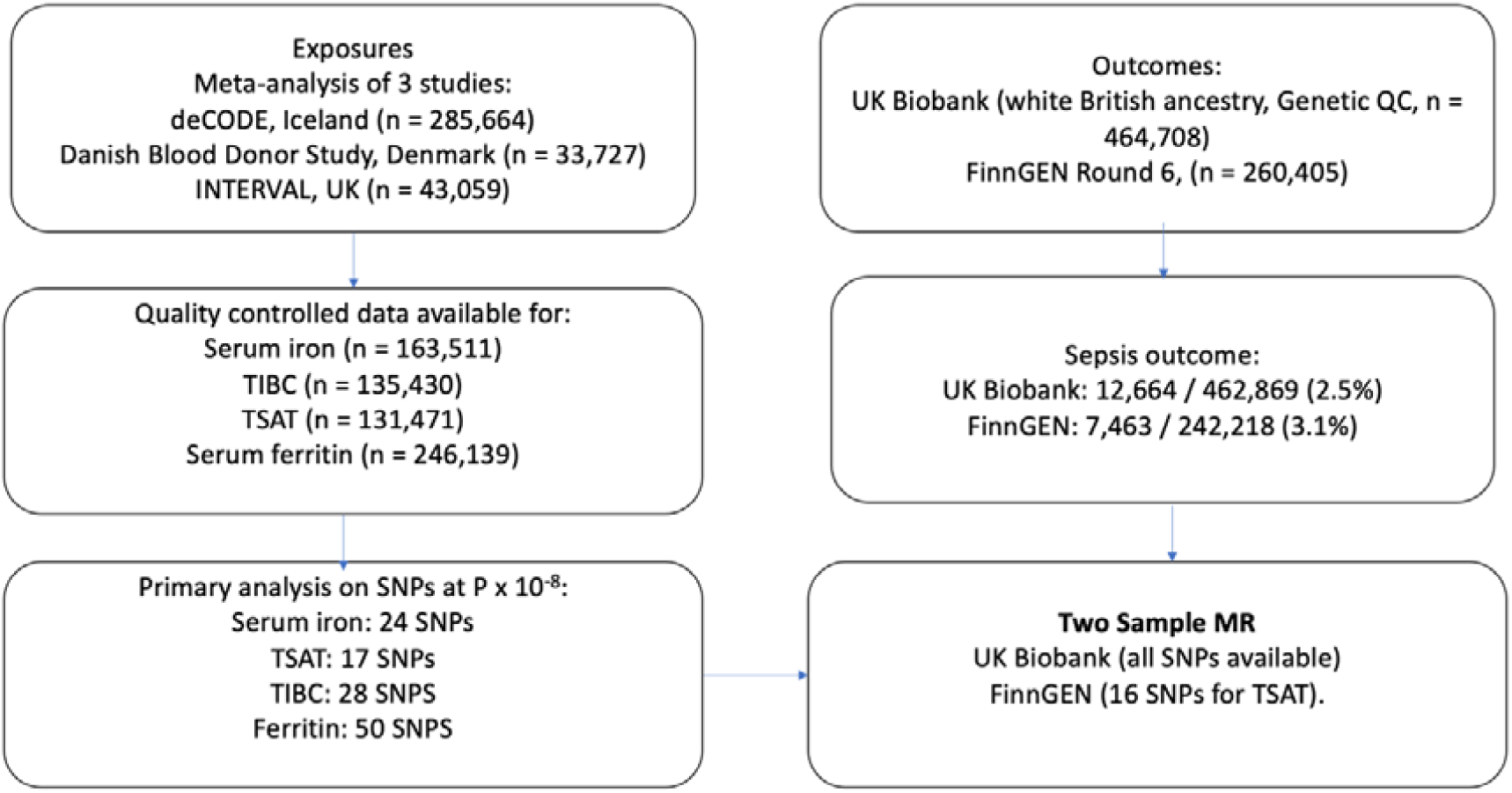
Flow chart of included studies, exposures, and outcomes.

##### Inverse variance weighted MR

We then went on to perform Two Sample Mendelian randomisation. We extracted SNPs from the outcome GWAS and harmonised effect alleles with the exposure SNPs, for each biomarker (Ferritin, Iron, TIBC, and TSAT) individually, and ran MR for each SNP on sepsis individually.^17^ To generate summary estimates for each biomarker, we performed inverse variance weighted (IVW) meta-analysis of each included SNP using the TwoSampleMR package in R.^21^

In IVW meta-analysis, increasing ferritin was associated with increased odds of sepsis (OR 1.17; 95% CI 1.04 – 1.29, for each SD increase in Ferritin, **Table 3**), with similar results for serum iron (OR 1.13; 95% CI 1.03 – 1.23), TSAT (OR 1.17 ; 95% CI 1.09 – 1.27), and with the opposite result for TIBC (OR 0.91; 95% CI 0.83 – 0.99), which would be expected as this marker decreases with increasing availability of iron. Scatter plots for TSAT – the biomarker with the strongest association is shown in **Figure 4**, while scatter plots for other biomarkers are available in **Supplementary Figure 6**.

**Figure 4:**
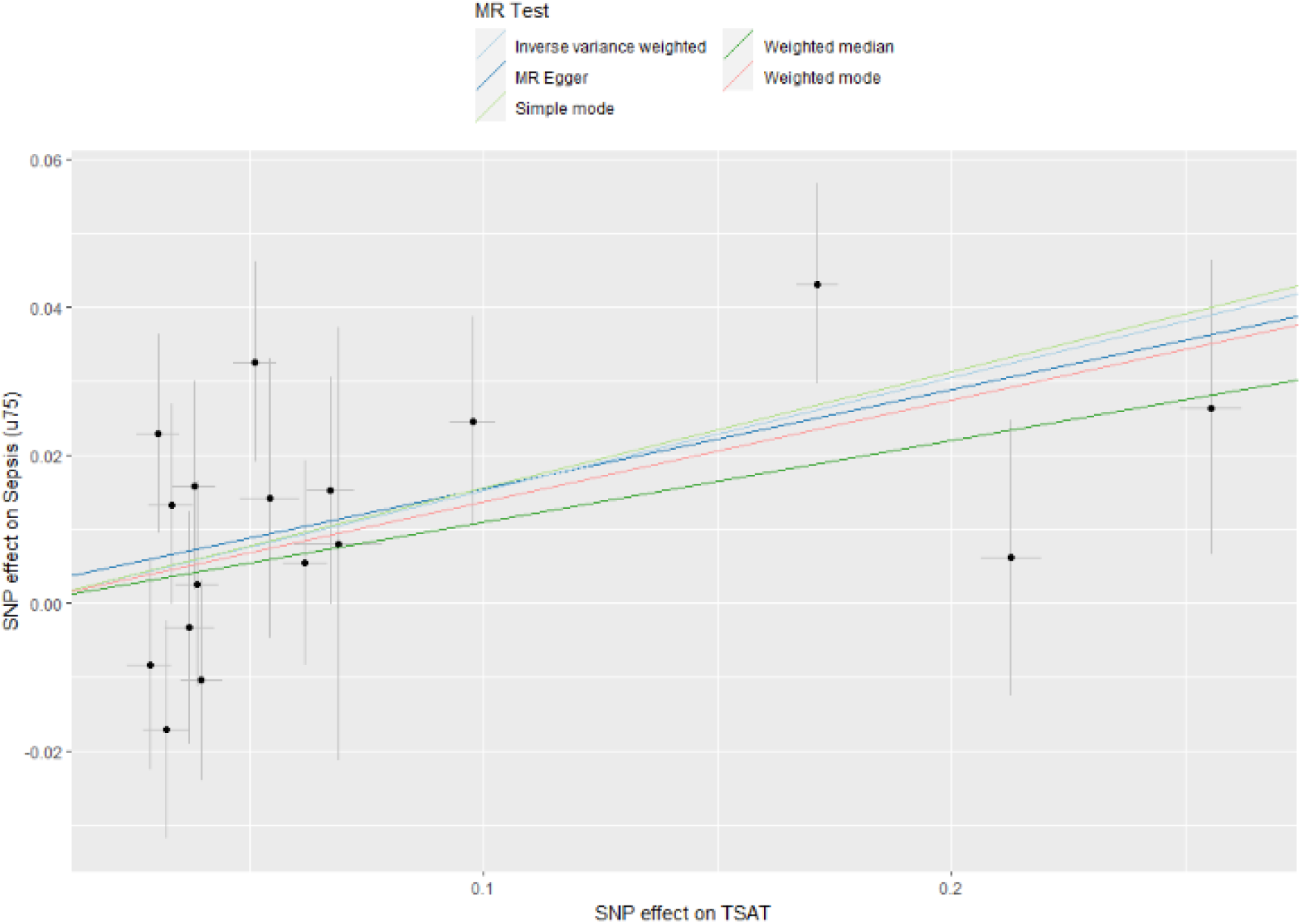
Scatter plot for increasing TSAT and risk of sepsis

##### Parallel analyses

We ran parallel analyses using outcomes derived in another cohort, FinnGen. FinnGen is a large, Finnish cohort study, with linked genetic and hospitalisation data. Details of the inclusion criteria are with the original study.^22^ Cases of sepsis were identified with the same ICD-10 coding as in UK Biobank, while details of the, study design, and GWAS methodology are in the methods and available at the FinnGen website.^22^ In FinnGen, there were 7,643 cases of sepsis and 234,755 controls, leading to a frequency of incident cases of 3.1%. As some SNPs were not available in this GWAS, we utilised LD proxies (r^2^ >0.8), if SNPS were not available.

In IVW meta-analysis, we identified broadly similar effect sizes for iron (OR 1.24; 95% CI 1.03 -1.45), although effect estimates for the other biomarkers were weaker, although all estimates were concordant. Scatter plots are available in **Supplementary Figure 7**.

We then went on to perform fixed effects meta-analysis across both cohorts. In this analysis, the summary estimates for each SD increase in iron biomarker was OR 1.09 (95% CI 1.02-1.15) for TSAT, OR 1.12 (95% CI 1.01 – 1.19) for iron, OR 1.08 (95% CI 0.98 – 1.18) for ferritin, and OR 0.93 (95% CI 0.93 – 0.98) for TIBC. These results are shown in **Figure 4** and **Table 3**.

**Figure 4:**
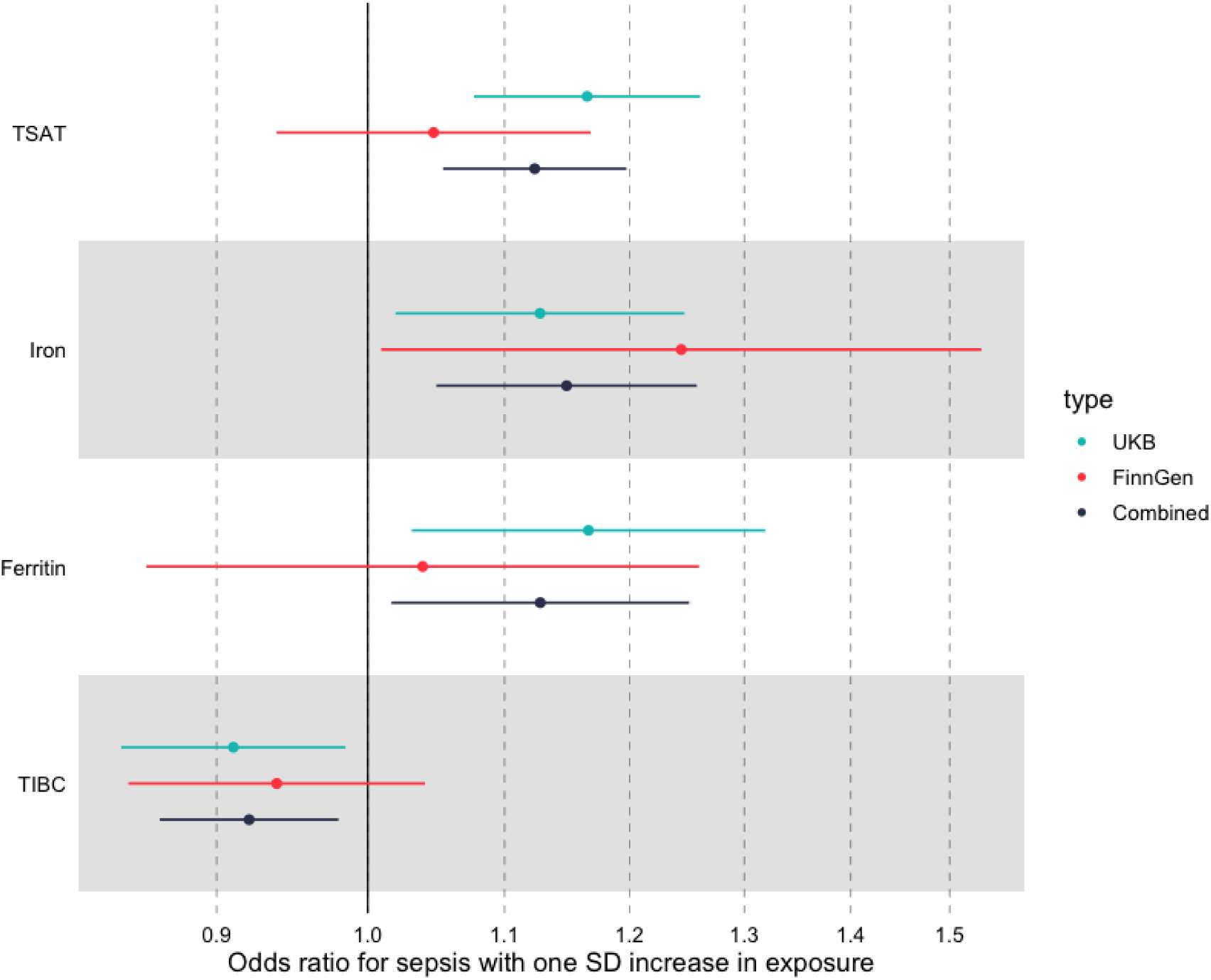
Summary IVW MR associations across both FinnGen and UK Biobank

##### Effect of measured iron status on estimates of risk

It is possible that the effect of iron related genetic variants varies by iron status. That is, it may be that increases in each biomarker have more or less of an effect in those who are clinically iron deficient or iron overloaded or those with and without anaemia. This analysis is important as the majority of iron supplementation occurs in those who either have biochemical iron deficiency (e.g. measured low ferritin), anaemia (e.g. low haemoglobin), or both. Initially, we compared estimates in those with or without anaemia on recruitment to UK Biobank.

We did this by performing stratified analyses within substrata of UK Biobank (e.g. those with or without anaemia) using a polygenic risk score (PRS) derived and weighted from the same variants used in the two sample MR.^23^ As we cannot reliably estimate the PRS-exposure association in UK Biobank, we cannot estimate a causal (MR) estimate using this approach, but can compare the effect sizes of the PRS in logistic regression in different strata. As we would not expect the PRS-exposure association to vary by exposure strata, this allows us to compare the effect size across two strata, and our effect sizes are reported per each change in standard deviation (SD) of the PRS, which was scaled to have a mean of 0 and an SD of 1.

In this approach, described in detail in the methods, strata are generated using the residuals of the regression of a PRS for each biomarker on haemoglobin (i.e. isolating a form of non-genetic variation in haemoglobin), in order to avoid potential bias possibly generated by stratification using a variable correlated with those in main analysis.^24^ Haemoglobin levels were available on admission to UK Biobank for nearly all European ancestry participants with genetic data (n = 448,978) and we stratified patients into anaemic and non-anaemic as per NICE definitions (Hb < 125g/L for women, Hb < 135g/L for men). Although the potential for collider bias was present, we found that each PRS had very little effect on haemoglobin levels, with the correlation between the original haemoglobin and the residuals of the regression of >0.99 in all cases.

For both anaemic and non-anaemic participants, we then performed logistic regression using our PRS. We identified – contrary to our expectations – that estimates of the odds of sepsis were generally higher in those with anaemia. For ferritin, the OR per one SD increase in the PRS for the anaemic cohort was 1.36 (95% CI 1.02 - 1.7), while it was only OR 1.13 (1.00 - 1.25) for the non-anaemic cohort. We identified similar effect changes for both TSAT and Iron (**Figure 5, Table 2)**, although effect estimates were similar for TIBC in both cohorts.

**Table 2:**
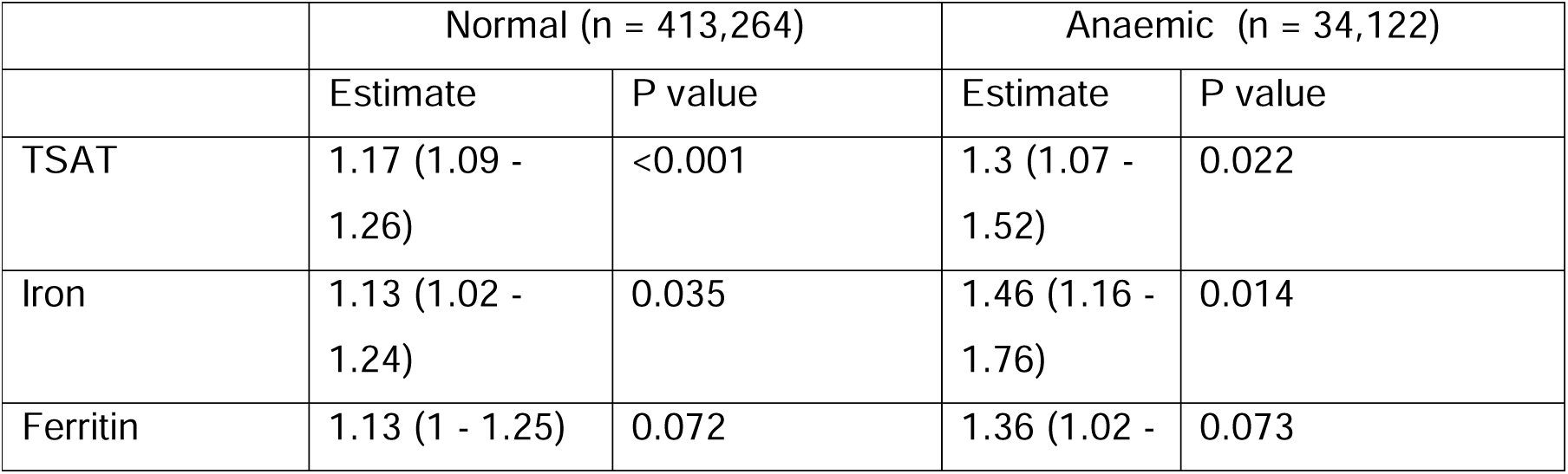
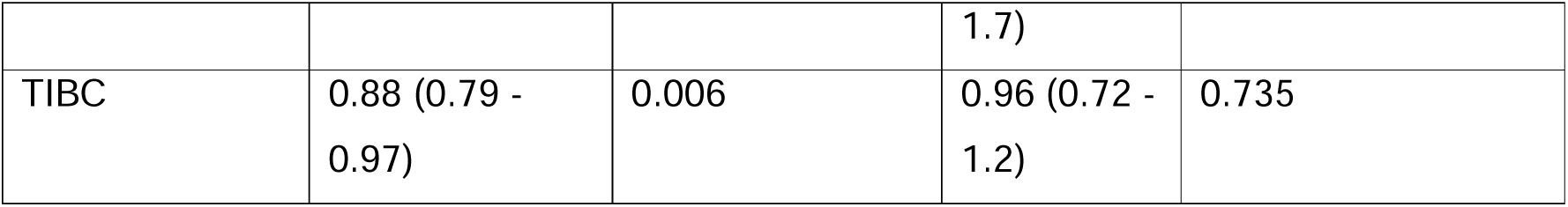
Odds ratios for each PRS in the anaemic and non-anaemic population in UK Biobank

**Table 3:** Meta-analysed IVW MR estimates for each biomarker across both FinnGen and UK Biobank.

|  | Odds ratio | Lower CI | Upper CI | P value |
| --- | --- | --- | --- | --- |
| *FinnGen* |  |  |  |  |
| TSAT | 1.05 | 0.94 | 1.16 | 0.41 |
| Iron | 1.24 | 1.03 | 1.45 | 0.04 |
| TIBC | 0.94 | 0.84 | 1.04 | 0.23 |
| Ferritin | 1.04 | 0.85 | 1.23 | 0.70 |
| *UK Biobank* |  |  |  |  |
| TSAT | 1.17 | 1.09 | 1.24 | <0.001 |
| Iron | 1.13 | 1.03 | 1.23 | 0.02 |
| TIBC | 0.91 | 0.83 | 0.99 | 0.02 |
| Ferritin | 1.17 | 1.04 | 1.29 | 0.01 |
| *Combined – IVW meta-analysis* |  |  |  |  |
| TSAT | 1.11 | 1.05 | 1.20 | <0.001 |
| Iron | 1.15 | 1.05 | 1.26 | 0.003 |
| TIBC | 0.92 | 0.87 | 0.98 | 0.01 |
| Ferritin | 1.13 | 1.02 | 1.25 | 0.02 |

**Table 3:** Odds ratios for a one SD increase in each PRS in each stratum of ferritin level

|  | Normal (50-450 ug/L) |  | Low Ferritin (<50 ug/L) |  | High ferritin (>450 ug/L) |  |
| --- | --- | --- | --- | --- | --- | --- |
|  | Estimate | P value | Estimate | P value | Estimate | P value |
| **TSAT** | 1.19 (0.97 - 1.42) | 0.128 | 1.33 (1.03 - 1.64) | 0.062 | 0.83 (0.29 - 1.37) | 0.498 |
| **Iron** | 1.31 (1.01 - 1.62) | 0.077 | 1.43 (1.02 - 1.85) | 0.084 | 0.41 (-0.3 - 1.13) | 0.016 |
| **Ferritin** | 1.19 (0.85 - 1.53) | 0.317 | 1 (0.52 - 1.47) | 0.985 | 0.52 (-0.34 - 1.37) | 0.130 |
| **TIBC** | 0.99 (0.75 - 1.23) | 0.956 | 0.92 (0.6 - 1.25) | 0.636 | 1 (0.45 - 1.55) | 0.989 |

**Figure 5:**
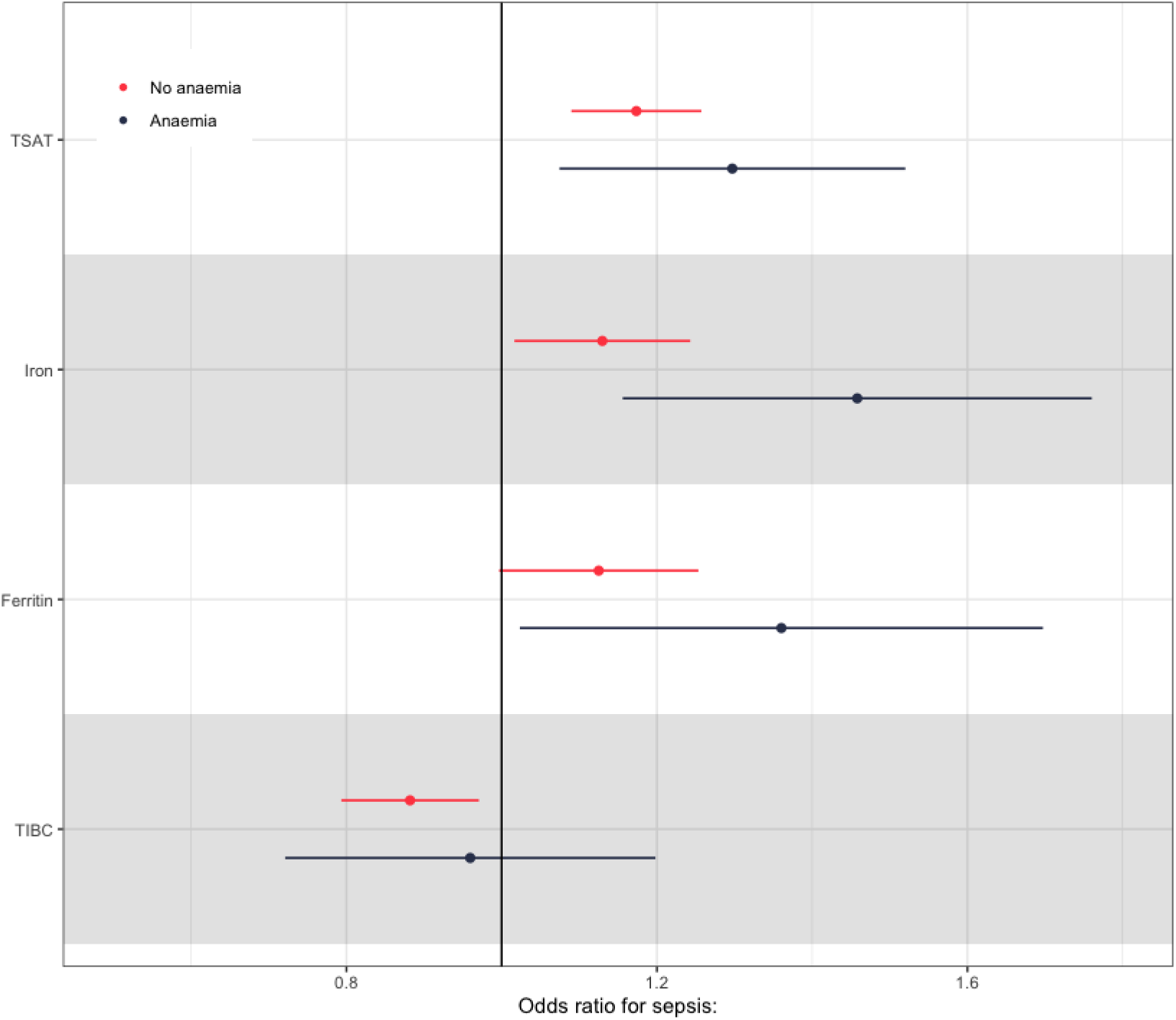
**Odds ratios for each SD increase in each PRS for sepsis in anaemic and non-anaemic population in UK Biobank**.

To explore this result further we then ran analyses in those with available ferritin levels from the observational analysis above (n = 75,277), in order to assess the impact of chronic iron stores on odds ratios generated by our PRS. As iron prescription is routinely indicated in those only with low ferritin, but occasionally given in those with higher levels, we chose to split this population into three strata using clinical definitions that are likely to affect the chance of a patient receiving treatment: iron deficient (<50ug/L), normal ferritin (50-450 ug/L), and high ferritin (>450 ug/L).

In this analysis, we generated PRS as for the haemoglobin analysis above, and used the samea analysis approach using residuals to generate strata to avoid collider bias.

For those with iron deficiency (low ferritin) and with normal ferritin, effect estimates were positive and similar (**Figure 6**), suggesting that increasing levels of iron biomarkers are associated with an increased rate of sepsis even in those with low/normal ferritin. In contrast, for the small number of participants with a ferritin of >450ug/L, effect estimates were reversed, although there was substantial uncertainty about the effect estimates due to a small sample size.

**Figure 6:**
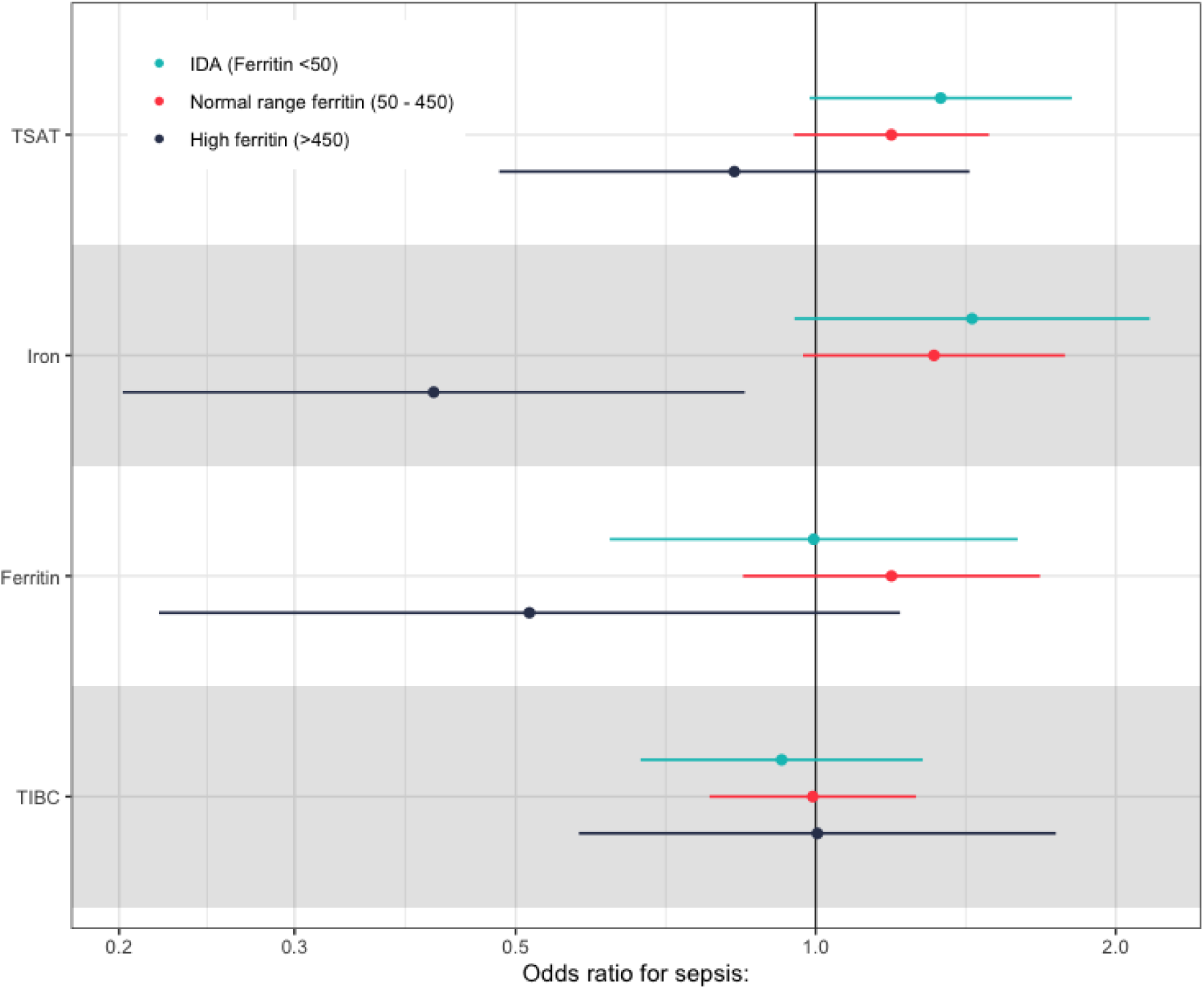
Odds ratios per unit SD increase in each PRS within each stratum of ferritin for each iron biomarker:

##### Testing assumptions of MR

We then tested the three main assumptions of MR and performed sensitivity analyses. Firstly, we tested for pleiotropy. Using our polygenic risk score (derived above) in all UK Biobank participants, we looked for associations with potential confounders of sepsis (**Supplementary Table S2)**. We found no strong evidence for association between age, sex and most clinical comorbidities, although there were associations identified for liver disease (OR 1.03; 95% CI 1.01-1.05, p = 0.002), and a history of cancer (OR 1.01; 95% CI 1.00-1.03, p = 0.01). Both are known associations of iron overload.

Subsequently, we performed a leave one out (LOO) analysis; whereby each SNP is removed from the MR one at a time, to ensure no individual SNP is driving the results. All results were robust to the LOO analysis, with little change in estimates (**Supplementary Figure 7**). Importantly, results were robust with or without inclusion of variants in the *HFE* gene, the commonest genetic cause of iron overload in the U.K.

Secondly, we meta-analysed the results using the MR-Egger approach and weighted median approach **(Figure 3, Supplementary Figure 5)**. Although the effect size of the estimates differed slightly, we found similar results regardless of statistical approach. Finally, we looked at the intercept of the MR-Egger analysis. If the intercept differs strongly from zero, this can represent confounding. However, all intercepts were close to 0, with formal tests for heterogeneity negative (Intercepts 0.002 for TSAT; -0.006 for iron; -0.001 for ferritin, -0.002 for TIBC, all p > 0.3).

##### Sensitivity analyses

As a sensitivity analysis for case definition, we ran the same analysis including cases of sepsis diagnosed in those of all ages (including those aged over 75). This showed broadly the same effect estimates as our primary analyses, although confidence intervals were broader. **Supplementary Table S3** includes these estimates, and the IVW weighted MA results with FinnGen.

## Discussion

In both observational and MR estimates, we found evidence of increasing odds of sepsis with increasing levels of iron and associated biomarkers. There was a suggestion that effect sizes were larger in those who were iron deficient or anaemic for both TSAT and iron, which may indicate that increases in iron even in those who have reduced iron stores (e.g. low ferritin) are associated with an increased risk of sepsis. Effect sizes were broadly similar in FinnGen, despite the well-recognised heterogeneity in sepsis presentation, definition, and coding across cohorts.^25^

One previous study has used two sample Mendelian randomisation to evaluate the relationship between iron biomarkers and sepsis, using a smaller cohort for identification of variants, and using a previously performed GWAS of sepsis in UK Biobank.^31^ In common with our study, they identified associations with iron biomarkers and the odds of sepsis (OR for iron = 1.21, 95% CI = 1.13–1.29, *p* = 3.16 × 10^−4^). However, this study included much fewer SNPs, as the GWAS used was around five to ten times smaller (n = 23,986 vs n = 246,139 for ferritin), meaning that only two SNPs were included for iron status, and three SNPS included for transferrin saturation and ferritin. This study was therefore heavily reliant on the inclusion of the rs1800562 SNP in *HFE*, which is the causative SNP for 85% of cases of hereditary haemochromatosis,^14^ making these results difficult to interpret outside this condition.

In addition, a further study performed an MR-PheWAS of iron levels in UK Biobank, with the strongest association being skin and soft tissue infection (OR 1.25; 95% CI 1.1-1.42 for each SD increase in serum iron).^14^ Although this study did not identify an association with sepsis, there were fewer cases (7,628 vs 15,614 in our analysis) at the time in UK Biobank. Secondly, a recent study of patients who carry mutations in the gene *HFE* leading to hereditary hemochromatosis, the most common cause of iron overload in the UK, identified an increased risk of pneumonia in male patients carrying *HFE* variants.^5^

In non-genetic literature, there have been a number of randomised trials of iron supplementation, some of which have measured infection as an outcome. These are summarised in a recent meta-analysis of 154 randomised trials of intravenous iron that found an increased risk of infection (RR 1.16; 95% CI 1.03-1.29), although there was marked heterogeneity across studies, and intravenous iron was compared against both “no iron”, and iron supplementation.^12^ As intravenous iron is associated with around a one SD increase in TSAT, our estimates are remarkable concordant with the trial data.

In complementary *in-vitro* work, one recent experimental study explored the effect of bacterial growth in serum taken from oral iron supplementation in healthy adults and identified an increase in TSAT from 42.1% (SD 12.5%) to 75.7% (SD 18%) and total serum iron increased from 30.3 μmol/L (SD 10.2) to 53.0μmol/L (SD 15.8) at four hours after iron supplementation, and identified TSAT as the key driver of bacterial growth, with positive associations at all levels of transferrin saturation.

### Strengths and weaknesses

The current work combines both traditional observational epidemiology with two sample MR using high quality genetic and phenotypic data, and the Hospital Episode Statistic (HES) based coding, but has limitations. Firstly, in observational analyses, it is clear that there are large differences in covariates across ferritin levels as a result of the sampling frame for available testing results, and as a result of the observational cohort. For instance, the relatively complex pattern of association, identified in other biomarkers (e.g. lymphocyte count, BMI^26,27^), with increased odds of incident sepsis cases in those with severe iron deficiency likely represents the increased risk associated with the disease driving severe iron deficiency (e.g. malignancy).

Secondly, common to all MR based studies, interpretation of an estimate for lifelong exposure to iron via germline genetic variation when assessed in other contexts, such as iron supplementation, requires justification. Iron exposure via supplementation is generally short lived, increases iron biomarkers over a short period (hours to days), and by a much larger amount than via most common SNPs. It is difficult to directly compare iron supplementation (individual events leading to short, time limited increases in iron levels) with lifelong increased iron exposure from genetic variants. However, the odds ratio for sepsis with each increasing standard deviation of TSAT from our meta-analysis of both cohorts (OR 1.11, 95% CI 1.05-1.20), should be put in the above context that oral supplementation increases TSAT by around 2-3 standard deviations over 4-8 hours, and has longer term effects^29,32^; while intravenous iron increases TSAT by around a standard deviation for at least three months in non-anaemic populations with chronic kidney disease.^33^

Thirdly, the heterogeneity of sepsis presents both a strength and a weakness. Sepsis is caused by a variety of bacteria, fungi, and viruses, only some of which scavenge free iron as a nutritional source (e.g. *S. aureus*, a common and important pathogen, uses largely heme-derived iron, and bacterial growth has been shown to be independent of serum iron levels^8^). As we were unable to access pathogen specific data, we could not identify pathogen-specific effects. This is compounded by the heterogeneity of clinical presentation and coding of sepsis, which has the effect of also reducing power significantly.^25,30^ In that context, we were reassured to see similar effect sizes across both UK Biobank and FinnGen, supporting our main findings and supporting the biological plausibility of the effect.

Finally, our approach in using residuals from a PRS of iron to develop strata in which to generate strata specific estimates allows us to make inference on whether the effect of increasing levels of iron biomarkers are likely to be the same in important subgroups (e.g. anaemic patients). However, the reversal of the effect estimate for increasing iron in those with high (>450 ug/L) ferritin was unexpected. Although it is possible this effect estimate is real, it may be that this represents an unaccounted-for collider bias (those who have high ferritin despite a low PRS for iron have a more sinister cause for their hyperferritinaemia). Alongside this, we make the assumption that the PRS-exposure association remains similar across all strata of iron and ferritin levels, although this assumption is also made in non-linear MR approaches.^23,24^

### Implications

Although these estimates support the randomised trial literature, ongoing research should focus on estimating absolute risks of severe infection across differing situations and with differing baseline iron levels. Further mechanistic understanding of how iron homeostasis setpoint changes are linked to the risk of sepsis, and the implications for iron management within infection, are warranted. Given that iron homeostasis is critical for both pathogens and hosts, it is important to understand whether these changes in iron homeostasis alter sepsis incidence via simply increasing iron availability to pathogens, or by an altered immune response to pathogens. Our findings and those from recent trial and *in-vitro* data, provide further evidence that iron state is a predictor of risk of severe infection, and also suggest caution in iron supplementation in areas without robust evidence of benefit.

## Conclusion

Observational data shows ferritin levels have a U-shaped relationship with sepsis, but the odds of sepsis increase even within the normal range of iron levels. Mendelian randomisation analyses show that physiologically relevant increases in iron biomarkers causally associated with an increased risk of sepsis, regardless of baseline iron status. These findings are in keeping with the hypothesis that iron supplementation increases the risk of severe infection and suggest further research into the implications of perturbing iron homeostasis.

## Methods

### Study design

An observational cohort study and two sample Mendelian randomisation analysis.

### Data sources

For both analyses we primarily used UK Biobank, a volunteer cohort of approximately 500,000 participants, that contains both genetic, physical, and biomarker information, and is linked to UK electronic health records. Recruitment started in 2006, ended in 2010, and participants continue to be followed up.^16^ Data for this study was extracted in October 2021. UK Biobank received ethical approval from the Research Ethics Committee (REC reference for UK Biobank is 11/NW/0382). This study was performed under application number 52643.

For our MR analysis, exposures sources and our parallel analyses in FinnGen are detailed below.

### Study design – observational cohort

#### Data sources and definitions

Approximately half of all participants in UK Biobank have associated primary care data (n = 222,081), including laboratory results. For our observational cohort, we extracted all serum ferritin levels recorded in primary care data linked to UK Biobank participants. We excluded extreme values (>10,000). The index ferritin level was taken as the first recorded test result after the date the participant was recruited to UK Biobank (between 2006-2010). For participants with multiple ferritin test results, only the index ferritin level was used. All participants with available, linked data were included.

We extracted demographic information and potential confounders from UK Biobank data: smoking status, alcohol usage, Townsend Deprivation Index, BMI, renal function, liver disease, presence of malignancy, and baseline inflammation (CRP on recruitment to UK Biobank). Variables were derived from UK Biobank questionnaire or algorithmic data. Included variables were all measured on recruitment and thus preceded ferritin testing and outcome ascertainment.

#### Statistical approach

The observational analysis used the raw ferritin level to estimate the risk of sepsis using logistic regression models adjusted for age, sex, UK Biobank recruitment centre, and the potential confounders listed above.

As very low ferritin (due to illness leading to iron deficiency) may alter the linearity of estimates, we also used restricted cubic spline modelling, using the *rms* package in R, and calculated specific estimates excluding those with likely iron deficiency (serum ferritin <50ug/L).^34^

### Study design – Mendelian Randomisation

#### Source of instruments

For our instruments, we extracted GWAS summary statistics from a recent meta-analysis of three separate GWA studies on four iron biomarkers (iron, transferrin saturation (TSAT), total iron binding capacity (TIBC), and ferritin).^17^ The original studies were performed in Iceland (deCODE genetics, n = 285,664), Denmark (Danish Blood Donor Study, n = 33,727), and the UK (Interval Study, n = 43,059). Details on each studies recruitment, follow up, GWAS and the meta-analytic methodology are included in their publication.^17^ Each biomarker was inverse rank-normalised transformed before GWAS, so the effect sizes are reported as per standard deviation change. For each biomarker, this genetic heritability (the proportion of variance explained by the SNPs alone) was estimated to be 16% for Iron, 22% TSAT, 25% for TIBC, and 30% for Ferritin.

#### Choice of primary instrument

For a primary biomarker of acute iron status we chose TSAT as this represents the biomarker most rapidly altered by iron supplementation, is the best predictor of haemoglobin response due to iron supplementation, and has the strongest prior relationship with infection outcomes, ^8,35,36^

#### Definition of instrument

For each biomarker, we extracted all SNPs that were available in the HRC-imputed UK Biobank genetic data and exposure data and met a threshold of p < 5 × 10^−8^, then performed LD-clumping to remove correlated variants (max R^2^ 0.01), using the TwoSampleMR package.^21^

17 SNPs for TSAT with a mean F-statistic of 325 were included. (Conventionally, scores of >10 are consistent with strong instruments).^18^ The minimum F-statistic included was 36.4. For serum iron, we included 23 SNPs (mean F statistic 186.7), for TIBC 26 SNPs (mean F statistic 214.3), and 50 SNPs for Ferritin (mean F statistic 86.7).

As not all SNPs were available in FinnGen, we used harmonised, proxy SNPs where LD proxied SNPs were available, resulting in 16 SNPs available for TSAT, 22 for Iron, 25 for TIBC and 50 for Ferritin.

As the original GWAS were performed using transformed data, the effect sizes are in relation to standard deviation increases in rank-normalised transformed values. In the DECODE Icelandic cohort, the standard deviation on untransformed data for TSAT was 11.6%, for serum iron 6 µmol/L, for ferritin 224ug/L, and for TIBC it was 12.6 µmol/L.

##### Exposure statistics

For our primary analysis, we included independent (R< 0.1) SNPs that met genome wide significance (P < 5 x10^−8^) in the recent iron meta-analysis.^17^ Details of included SNPs, F-statistics, and data on the other biomarkers is included in **Supplement S4**.

##### Outcome definition

Cases of sepsis were identified in UK Biobank linked ICD-coded hospital admission data. ICD-10 codes A02, A39, A40 and A41 were used to identify sepsis. Cases were included if the code was in either the primary or secondary diagnostic position in the linked Hospital Episode Statistic (HES) data. All patients who had a code for sepsis before the age of 75 were included (13,260/16,173, or 82% of total cases), as the relative contribution of germline genetic variation on iron status on sepsis in extreme age was expected to be much smaller. As a sensitivity analysis, we included cases of sepsis at all ages.

##### GWAS

A case-control GWAS was performed of using regenie v2.2.4, on all UK Biobank participants of European ancestry.^20^ In house algorithms were used to perform quality control and define ancestry with details available elsewhere.^19,36^ GWAS was performed adjusting for age, sex, genetic chip, UK Biobank assessment centre and the first ten principal components.^19^

Full details on the methodology are available elsewhere, but are described below in brief.^19^ regenie was chosen as it reduces type 1 error when there is significant case control imbalance, as compared to other methods such as BOLT-LMM.^20^ Manhattan and QQ-plots for GWAS performed as part of this study are available in **Supplementary Figure 4 and 5**. Summary statistics are available at the IEU Open GWAS repository (https://gwas.mrcieu.ac.uk/datasets/ieu-b-5066/). The top GWAS hits per chromosome are described in **Supplement S3**.

##### Genotyping and imputation

The data release for this analysis contains the cohort of successfully genotyped samples (n=488,377). 49,979 individuals were genotyped using the UK BiLEVE array and 438,398 using the UK Biobank axiom array. Pre-imputation QC, phasing and imputation are described elsewhere.^23^ In brief, prior to phasing, multiallelic SNPs or those with MAF ≤1% were removed. Phasing of genotype data was performed using a modified version of the SHAPEIT2 algorithm. Genotype imputation to a reference set combining the UK10K haplotype and HRC reference panels was performed using IMPUTE2 algorithms.

The analyses presented here were restricted to autosomal variants using a graded filtering with varying imputation quality for different allele frequency ranges. Therefore, rarer genetic variants are required to have a higher imputation INFO score (Info>0.3 for MAF >3%; Info>0.6 for MAF 1-3%; Info>0.8 for MAF 0.5-1%; Info>0.9 for MAF 0.1-0.5%) with MAF and Info scores having been recalculated on an in-house derived ‘European’ subset.^23^

##### Data quality control

Individuals with sex-mismatch (derived by comparing genetic sex and reported sex) or individuals with sex-chromosome aneuploidy were excluded from the analysis (n=814).

##### Ancestry

We restricted the sample to individuals of ‘European’ ancestry as defined by an in-house kmeans cluster analysis performed using the first 4 principal components provided by UK Biobank in the statistical software environment R. The current analysis includes the largest cluster from this analysis (n=464,708).^23^

### Statistical approach

Univariable two sample, Mendelian randomisation (MR) analyses were conducted using the R package *TwoSampleMR* to investigate total effect estimates between genetically predicted iron status (for each biomarker individually) with sepsis.^21^ This was based on the inverse variance weighted (IVW) method, which estimates the causal effect of an exposure on an outcome by combining ratio estimates using each variant in a fixed effect meta-analysis.

### Testing for interactions with respect to ferritin and haemoglobin levels

As total iron state (e.g. iron deficiency defined by low ferritin) might affect MR estimates for the acute iron biomarkers, we ran specific analyses in those with varying levels of Hb and ferritin.

To do this, we generated weighted allelic polygenic risk score (PRS) generated for each UK Biobank participant for each biomarker. All SNPs from the MR analysis were included, with weighting of each SNP by the effect size of that SNP. With certain underlying assumptions, this allelic risk score can be then used to perform MR.^38^ We then extracted haemoglobin levels (from blood taken on recruitment to UK Biobank), and ferritin levels (from the primary care data, as detailed above). We also extracted sepsis cases occurring after blood sampling, using the same approach as in the observational data.

We then performed ordinary least-squares (OLS) regression for each PRS on haemoglobin and ferritin, individually, and extracted the residuals from this regression. In effect, this removes the PRS (“genetic”) effect on each biomarker, leaving only the non-genetic components, an approach previously used to assess the dose-response relationship of other biomarkers.^13,24,39^ We then stratified these residuals. For haemoglobin, we stratified into anaemic and non-anaemic (Hb < 125g/L for women, Hb < 135g/L for men). For ferritin, we split into three cohorts: Iron deficient (<50ug/L), normal ferritin (50-450 ug/L), and high ferritin (>450 ug/L). This use of residuals for strata is required as a naïve analysis stratifying on raw data would be potentially biased, as the PRS will associate with each strata (e.g. those with lower PRS for iron more are likely to be anaemic).

Subsequently, within the generated strata, we ran a logistic regression for each PRS on sepsis cases. In effect, this allows us to estimate whether MR estimates are altered in those who have differing levels of haemoglobin and/or ferritin.

### Sensitivity analyses

Mendelian randomization analysis assumes that a genetic instrument used to proxy a risk factor (1) is associated with the risk factor (“relevance”), (2) does not share a common cause with the outcome (“exchangeability”), and (3) affects the outcome only through the risk factor (“exclusion restriction”). To test relevance, we measured the F-statistic for each exposure for each biomarker, while the previous meta-analysis has shown a reasonable proportion of variance explained (>15% for each biomarker).^17^

To test the assumption of exclusion restriction we tested a weighted allelic risk score (described below) against confounders (sex, BMI, diabetes, deprivation and, smoking status) by OLS regression using a specific MR-PheWAS software (PHESANT).^40^ Finally we also performed a set of statistical sensitivity analyses that test assumptions of MR, including different meta-analytic methods: MR-Egger, weighted median, weighted mode, and iterative leave one out analyses, all of which test various assumptions of the MR approach.

#### Parallel analyses

The linear analysis was run in parallel using the FinnGen R6 summary statistics for sepsis.^22^ As some SNPs were not available in this cohort, SNP proxies (LD > 0.8) were selected and harmonised to ensure the effect alleles matched using the *gwasVCF* and *TwoSampleMR* package if a SNP was not available.^21^

We report both the FinnGen results, and a fixed effects meta-analysis of both cohorts using the *meta* package in R. ^41^

#### Power calculation

Formally, we calculated power with a sample size of 460,000 participants, an R^2^ of 0.20, alpha at 0.05 and a case-to-control ratio of 40 to be 99.4% to detect an OR of 1.1 or more using the approach of Burgesss et al.^42^

### Reporting

This study is reported in line with the STROBE-MR reporting guidelines, available in the supplement.^43^

### Code and data availability

Access to individual data for UK Biobank is via application to the independent data access committee. For this paper, summary outcome GWAS were released to the OpenGWAS repository (https://gwas.mrcieu.ac.uk/datasets/ieu-b-5066/), and summary exposure data are available at deCODE.genetics website (https://www.decode.com/summarydata/), and therefore all MR results can be replicated using the *TwoSampleMR* package.^21^

## Data Availability

Access to individual data for UK Biobank is via application to the independent data access committee. For this paper, summary outcome GWAS are available at the OpenGWAS repository (https://gwas.mrcieu.ac.uk/datasets/ieu-b-5066/), and summary exposure data are available at deCODE.genetics website (https://www.decode.com/summarydata/), and therefore MR results can be replicated using the TwoSampleMR package.
 

