## Supplementary material for "Causal associations between iron status and sepsis: a Mendelian randomisation analysis": S1 Figs and tables (from another version of this preprint: version 1, 2022-04-29)

**Figure S1: Flow of participants through the observational study**
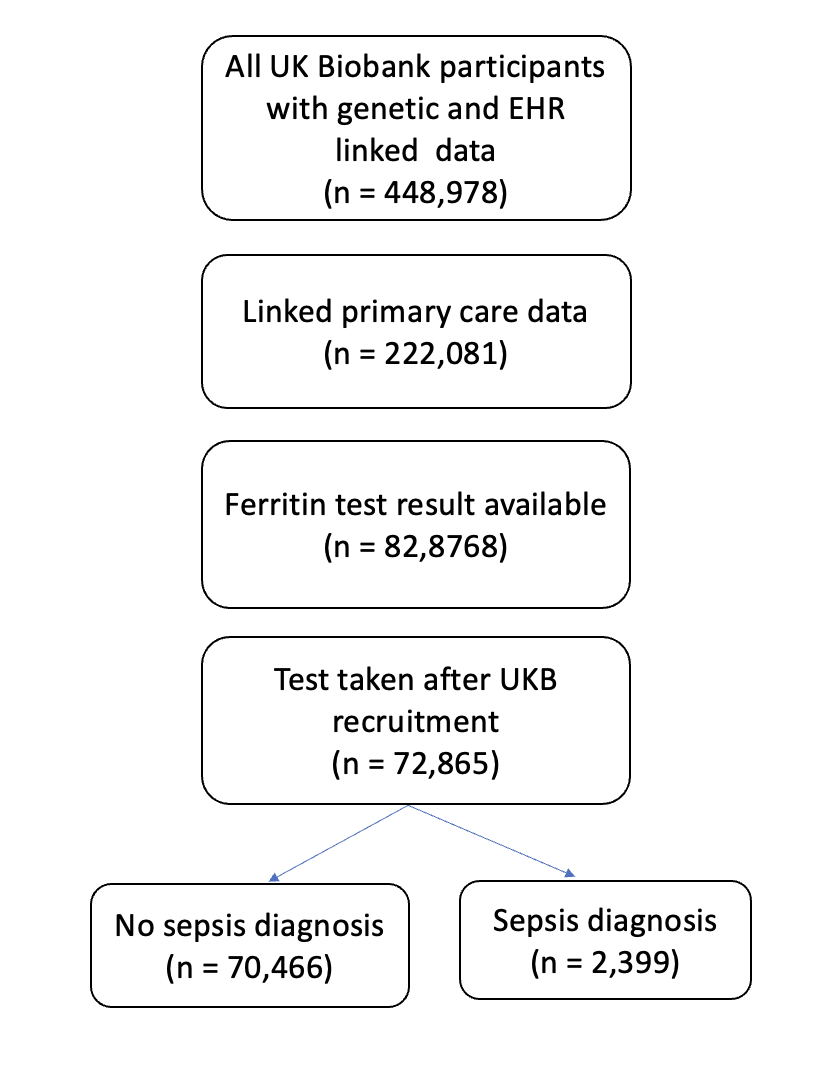

**Figure S2: Histogram of Ferritin (log-normalised) for men and women (all tests)**

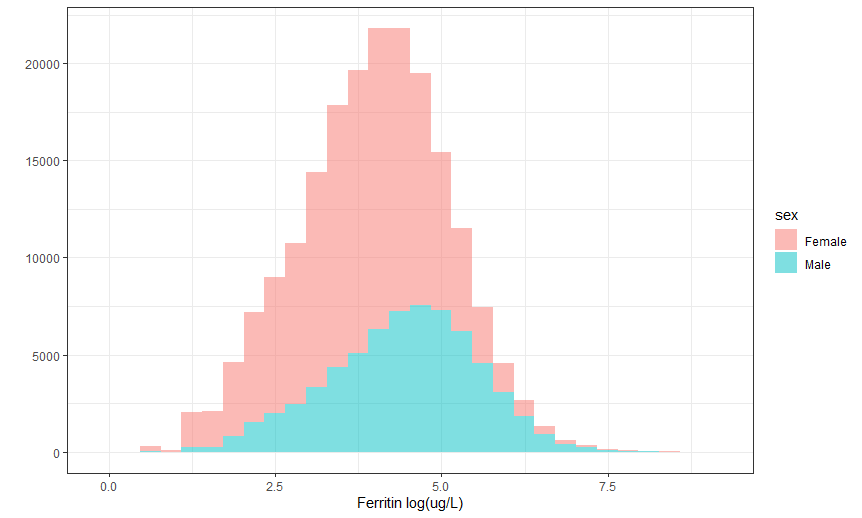

Supplementary Table 1: Regression outputs for ferritin. Biobank recruitment centre removed for brevity.

| term | estimate | lower | upper | p.value |
| --- | --- | --- | --- | --- |
| (Intercept) | 0.000 | −1.524 | 1.525 | 0.000 |
| value_100 | 1.050 | 1.036 | 1.064 | 0.000 |
| age_at_entry | 1.065 | 1.059 | 1.071 | 0.000 |
| sexMale | 1.773 | 1.685 | 1.861 | 0.000 |
| bmi | 1.054 | 1.046 | 1.061 | 0.000 |
| liver_disease | 1.503 | 1.274 | 1.731 | 0.000 |
| cancer | 1.892 | 1.783 | 2.001 | 0.000 |
| townsend | 1.036 | 1.022 | 1.050 | 0.000 |
| alcohol_intake_freq.L | 0.876 | 0.454 | 1.297 | 0.537 |
| alcohol_intake_freq.Q | 2.024 | 1.618 | 2.431 | 0.001 |
| alcohol_intake_freq.C | 0.772 | 0.459 | 1.085 | 0.105 |
| alcohol_intake_freq^4 | 1.139 | 0.939 | 1.339 | 0.201 |
| alcohol_intake_freq^5 | 0.902 | 0.766 | 1.038 | 0.136 |
| alcohol_intake_freq^6 | 0.930 | 0.823 | 1.038 | 0.188 |
| as.factor(smoking).L | 0.895 | 0.593 | 1.197 | 0.471 |
| as.factor(smoking).Q | 1.618 | 1.388 | 1.848 | 0.000 |
| as.factor(smoking).C | 0.831 | 0.712 | 0.949 | 0.002 |

**Supplementary Figure 3: Restricted cubic spline models of the association between ferritin and sepsis in both men and women. A: Men, B, Women**

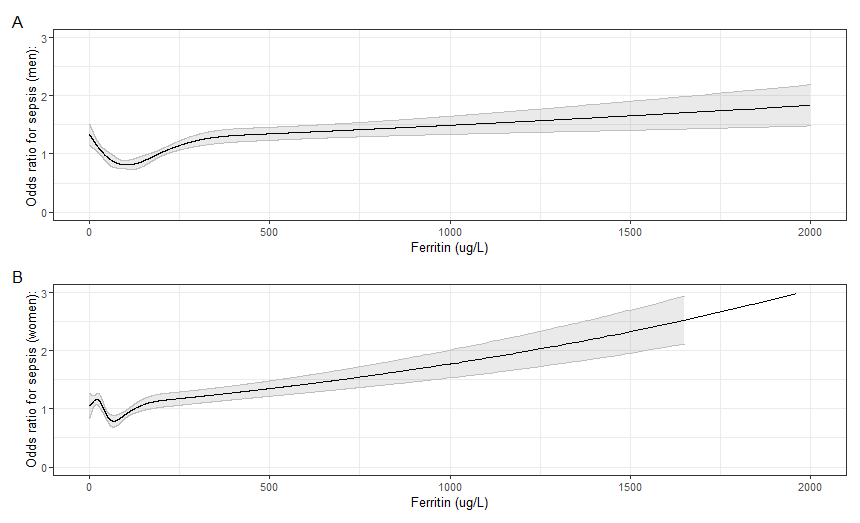

**Supplementary Figure 4: GWAS and QQ plots for included GWAS.**

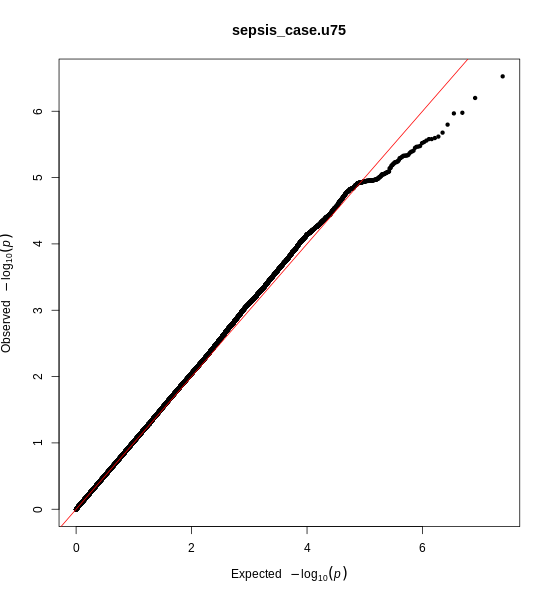

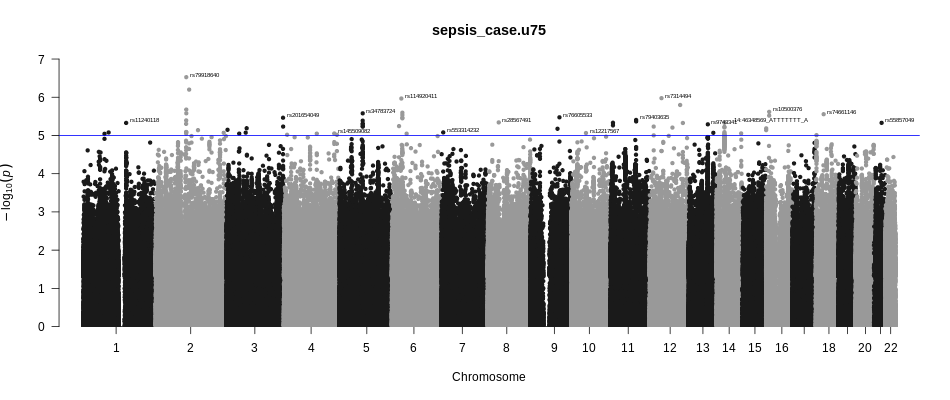

**Supplementary Figure 5: Scatter plots for all four biomarkers on sepsis (A: TSAT, B: Iron, C: Ferritin, D: TIBC)**

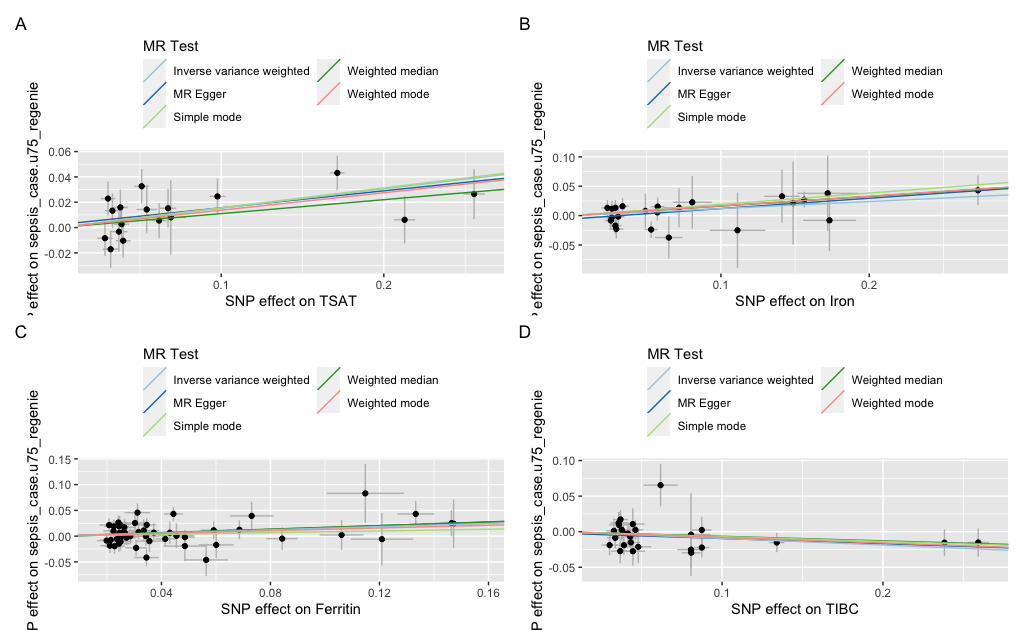

**Supplementary Figure 6: Scatter plots for FinnGen data (A: TSAT, B: Iron, C: Ferritin, D: TIBC)**

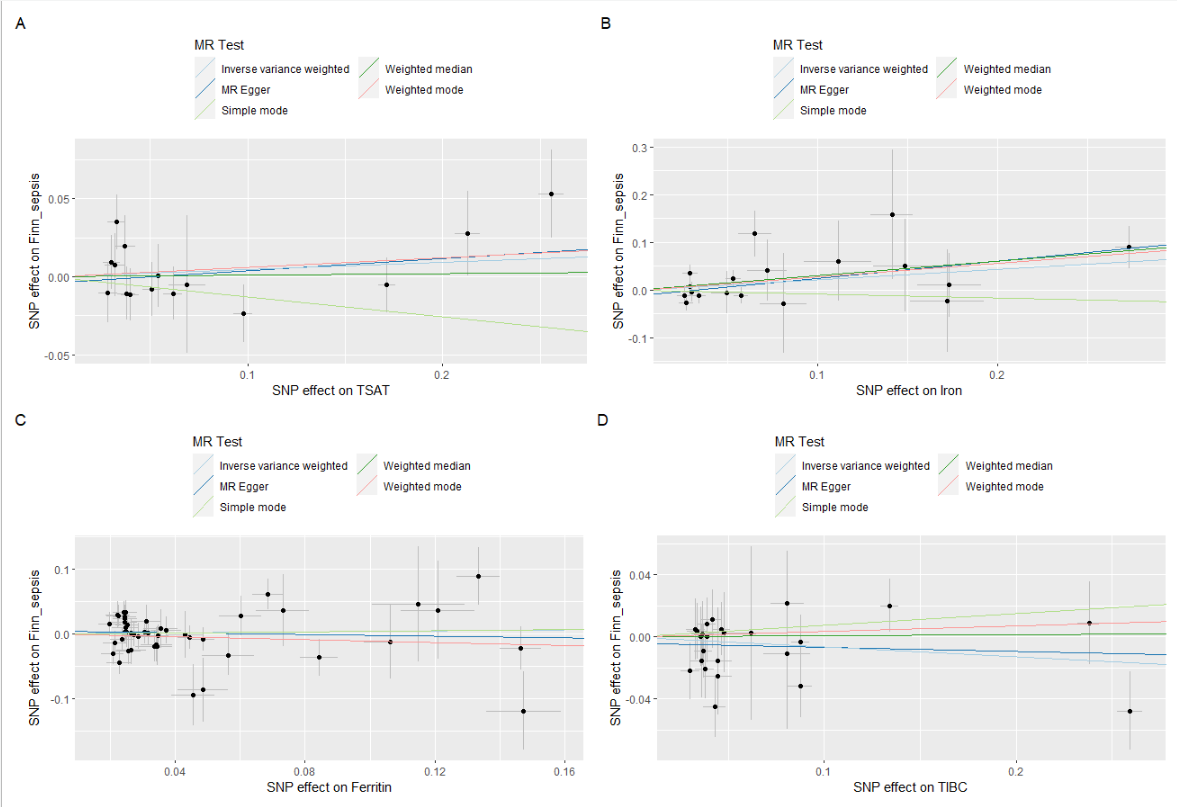

**Table S1: Associations between allelic risk score and potential confounders in UK Biobank**

| method | beta | lower | upper | pvalue | n |
| --- | --- | --- | --- | --- | --- |
| Alcohol intake frequency. |  |  |  |  |  |
| Ferritin | 1.008 | 1.003 | 1.013 | 0.003 | 462161 |
| Iron | 1.004 | 0.999 | 1.009 | 0.114 | 462161 |
| TIBC | 1.009 | 1.003 | 1.014 | 0.001 | 462161 |
| TSAT | 1.003 | 0.997 | 1.008 | 0.328 | 462161 |
| Date K76 first reported (other diseases of liver) |  |  |  |  |  |
| Ferritin | 1.019 | 1.001 | 1.038 | 0.037 | 450519/12304(462823) |
| Iron | 1.027 | 1.009 | 1.046 | 0.003 | 450519/12304(462823) |
| TIBC | 0.982 | 0.965 | 1.000 | 0.046 | 450519/12304(462823) |
| TSAT | 1.029 | 1.010 | 1.047 | 0.002 | 450519/12304(462823) |
| Diabetes diagnosed by doctor |  |  |  |  |  |
| Ferritin | 1.013 | 1.000 | 1.027 | 0.057 | 439061/22331(461392) |
| Iron | 1.006 | 0.992 | 1.019 | 0.414 | 439061/22331(461392) |
| TIBC | 1.002 | 0.989 | 1.016 | 0.768 | 439061/22331(461392) |
| TSAT | 1.008 | 0.994 | 1.021 | 0.269 | 439061/22331(461392) |
| Body mass index (BMI) |  |  |  |  |  |
| Ferritin | 0.997 | 0.994 | 1.000 | 0.063 | 461274 |
| Iron | 0.996 | 0.993 | 0.998 | 0.002 | 461274 |
| TIBC | 1.004 | 1.001 | 1.007 | 0.003 | 461274 |
| TSAT | 0.999 | 0.996 | 1.002 | 0.450 | 461274 |
| Date N18 first reported (chronic renal failure) |  |  |  |  |  |
| Ferritin | 0.993 | 0.980 | 1.006 | 0.283 | 439557/23266(462823) |
| Iron | 0.993 | 0.979 | 1.006 | 0.272 | 439557/23266(462823) |
| TIBC | 1.006 | 0.993 | 1.019 | 0.396 | 439557/23266(462823) |
| TSAT | 0.999 | 0.986 | 1.013 | 0.920 | 439557/23266(462823) |
| Cancer diagnosed by doctor |  |  |  |  |  |
| Ferritin | 1.004 | 0.993 | 1.015 | 0.494 | 424971/36155(461126) |
| Iron | 1.011 | 1.000 | 1.022 | 0.046 | 424971/36155(461126) |
| TIBC | 0.994 | 0.983 | 1.005 | 0.278 | 424971/36155(461126) |
| TSAT | 1.014 | 1.003 | 1.025 | 0.010 | 424971/36155(461126) |
| Age at death |  |  |  |  |  |
| Ferritin | 0.998 | 0.987 | 1.009 | 0.669 | 32497 |
| Iron | 1.004 | 0.993 | 1.015 | 0.447 | 32497 |
| TIBC | 1.003 | 0.993 | 1.014 | 0.526 | 32497 |
| Townsend deprivation index at recruitment |  |  |  |  |  |
| Ferritin | 1.000 | 0.998 | 1.003 | 0.774 | 462279 |
| Iron | 0.998 | 0.995 | 1.001 | 0.160 | 462279 |
| TIBC | 0.999 | 0.997 | 1.002 | 0.686 | 462279 |
| TSAT | 0.999 | 0.996 | 1.002 | 0.556 | 462279 |
| Pack years of smoking |  |  |  |  |  |
| Ferritin | 1.001 | 0.995 | 1.006 | 0.803 | 143129 |
| Iron | 1.001 | 0.996 | 1.007 | 0.621 | 143129 |
| TIBC | 0.998 | 0.993 | 1.003 | 0.387 | 143129 |
| TSAT | 1.003 | 0.998 | 1.008 | 0.277 | 143129 |

**Supplementary Figure S7: MR Leave one out analysis**

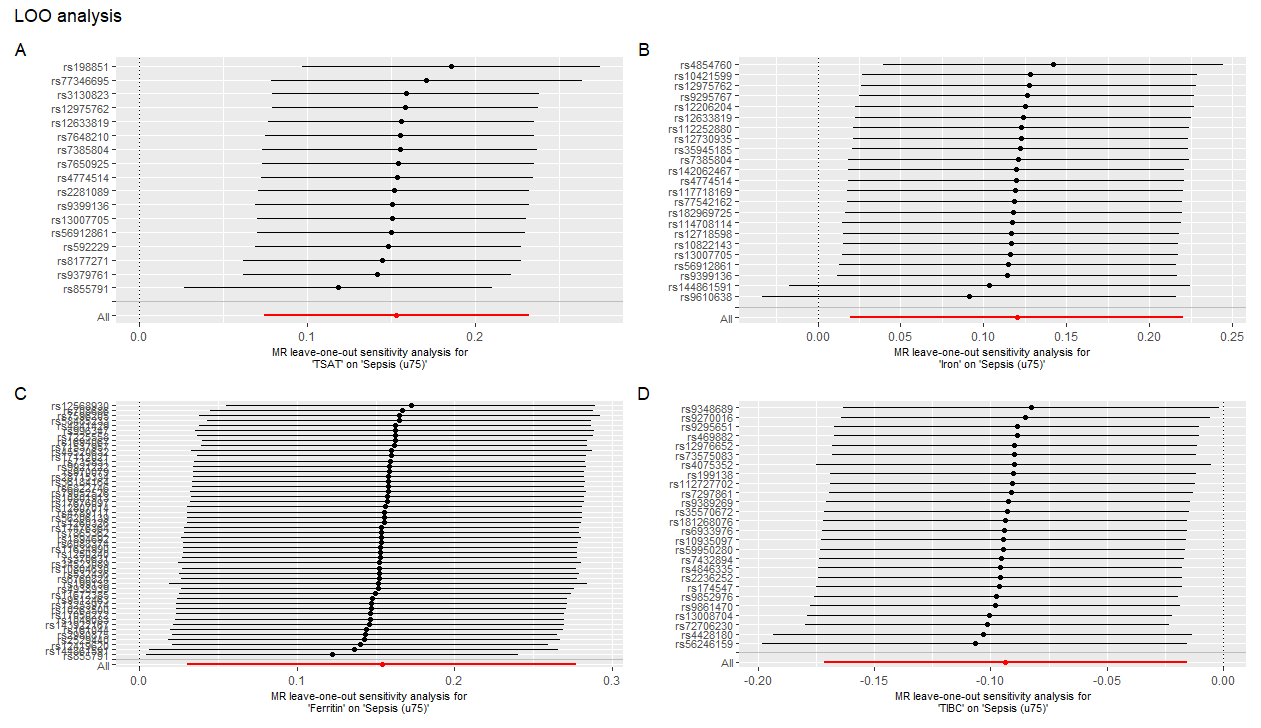

**Supplementary Figure S6: Scatter plots for the TSAT, serum iron, serum ferritin, and TIBC on all cases of sepsis (sensitivity analysis including cases >75 years, A: TSAT, B: Iron, C: Ferritin, D: TIBC)**

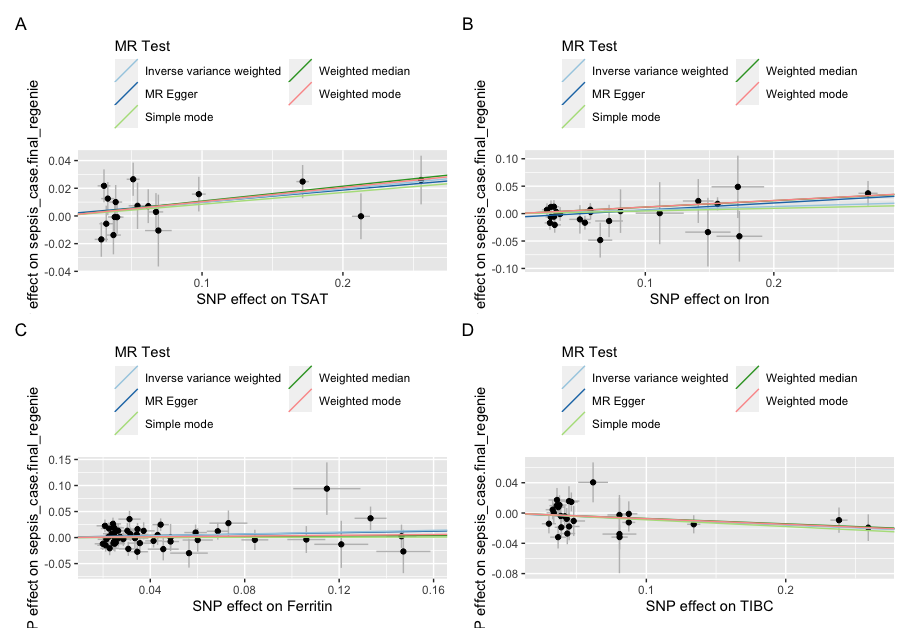

**Supplementary Table S2: All case analysis using UK Biobank**

|  | Odds ratio | Lower CI | Upper CI | P value |
| --- | --- | --- | --- | --- |
| *UK Biobank (all cases)* | | | | |
| Iron | 1.07 | 0.98 | 1.16 | 0.16 |
| TIBC | 0.92 | 0.86 | 0.99 | 0.03 |
| TSAT | 1.10 | 1.03 | 1.17 | 0.01 |
| Ferritin | 1.09 | 0.99 | 1.20 | 0.11 |
| *Combined – IVW meta-analysis* | | | | |
| Iron | 1.12 | 1.01 | 1.19 | 0.04 |
| TIBC | 0.93 | 0.88 | 0.98 | 0.01 |
| TSAT | 1.09 | 1.02 | 1.15 | 0.01 |
| Ferritin | 1.08 | 0.98 | 1.18 | 0.11 |
